# Facilitators and barriers to dietary choices among older adults living in rural Edo, South-south, Nigeria

**DOI:** 10.64898/2026.03.16.26348208

**Authors:** Onosolesena D. Idiakheua, Elizabeth A. Williams, Ohilebo A. Abass, Ebabhiele J. Idiakhua, Viren Ranawana, Robert Akparibo

**Author notes:** Corresponding author: Onosolesena D. Idiakheua.

## Abstract

**Background:** Population ageing is accelerating, with the fastest growth occurring in low-and middle-income countries. Adequate nutrition is central to healthy ageing, yet little is known about the factors shaping dietary behaviours among older adults in rural African settings, where structural constraints may strongly influence dietary choice. This study explored the facilitators and barriers influencing the dietary choices among older adults aged 60 years and above living in rural communities of Edo Central, Nigeria.

**Methods:** This exploratory qualitative study was guided by the Socio-Ecological Model (SEM). Semi-structured, in-depth interviews were conducted with 22 older adults. Interview transcripts were analysed thematically using NVivo 14, with findings mapped across individual, environmental, community, and policy/system levels of the SEM.

**Results:** Fourteen subthemes were identified and organised into four overacting SEM domains. Individual drivers identified included a deep knowledge of nutrient-rich diets and a preference for natural, minimally processed foods, as well as community drivers, including cultural and traditional norms and market access, which were the facilitators. Environmental drivers, including physical and economic access, and policy/system drivers, including government policies/subsidies, food prices and inflation, were identified as the main barriers.

**Conclusion:** Dietary choices among older adults in rural Nigeria are shaped predominantly by structural and food-system constraints rather than by individual knowledge alone. Policies aimed at improving nutrition in ageing populations should prioritise strengthening rural food systems, supporting smallholder agriculture, stabilising food prices, and developing targeted social protection programmes for older adults.

## Introduction

The global population of older adults, defined as individuals aged 65 years and above (1), is undergoing rapid growth, projected to reach 2.2 billion by the 2070s, with Sub-Saharan Africa, particularly Nigeria (estimated 7.24 million), making a significant contribution (2). Promoting active and independent ageing is an important public health priority that relies heavily on healthy lifestyle factors (3). Dietary choices and nutrition play a vital role in successful ageing, as a balanced diet is recognised as a key modifiable factor for lowering the risk and slowing the progression of common chronic diseases, preventing infection and reducing the risk of frailty (4, 5). These nutritional issues (e.g., double burden malnutrition, sarcopenia, and ageing anorexia) are compounded by the ongoing epidemiological transition in Nigeria, where non-communicable diseases (NCDs) are increasingly prevalent, and mortality rates are rising, making preventative nutritional strategies a priority (6). Determinants of healthy eating are classified across individual (physiological, psychological), interpersonal (social, cultural), environmental, and policy factors (7), all of which contribute to achieving optimal nutrient intake and physical function of older adults (8, 9).

Despite the consensus that multiple interconnected factors facilitate or hinder healthy dietary behaviour among older adults (10), research is limited in low- and middle-income countries, especially in Africa. Although some studies in Nigeria have quantitatively examined specific elements like fruit and vegetable intake among older adults (11, 12), qualitative evidence to help better understand and appreciate older adults’ dietary practices or food behaviours is lacking. The only two existing studies tend to focus narrowly on specific nutrient selection behaviours, underscoring the limited qualitative, in-depth information available on the holistic experience of dietary choices, diet patterns and overall eating behaviours in this population. Older adults in Nigeria, including those living in the Edo Central District, face unique socioeconomic, structural, and environmental constraints (including limited infrastructure and markets) that restrict access to nutritious food and are underrepresented in the existing literature.

To address this knowledge gap, this is the first study adopted a qualitative design to identify the facilitators and barriers to dietary choices, and practices/behaviours among older adults living in rural areas, Edo State, Nigeria. The findings provides context-specific evidence to support the development of targeted public health interventions to promote adequate and balanced nutrition for older adults in Nigeria.

## Methods

### Study Design and Setting

This qualitative study was conducted in the Edo Central Senatorial District of Edo State, Nigeria. The district has a population of 1,015,894, with older adults comprising 3.1% of the population (13). Edo Central is in the South-South geopolitical zone (lying roughly between longitudes 6°04’E and 6°43’E and latitudes 5°44’N and 7°34’N) and includes five Local Government Areas (LGAs): Esan Central, Esan North-East, Esan South-East, Esan West, and Igueben (14). The district is predominantly rural, characterised by a strong Esan ethnic identity and livelihood patterns focused on subsistence agriculture (cash crops include oil palm, cassava, and yams) and petty trading (15). The dependence on agriculture and informal trading directly influences the economic security and food access of the rural older adult population, making this a highly relevant setting to explore nutritional determinants.

#### Study Participants and Sampling

The study focused on older adults aged 60 years and older living in rural communities within the Edo Central Senatorial District. This population was selected due to their heightened vulnerability to nutritional risk, which stems from limited income, physical decline, health issues, and restricted access to diverse foods and healthcare services compared to urban counterparts (16, 17). Purposive sampling was employed to intentionally select participants capable of providing rich and relevant insights (18). Sampling was guided by diversity criteria, including gender, socioeconomic status (e.g. retired civil servants, subsistence farmers, petty traders), and assumed differences in dietary habits, to ensure a wide range of experiences. The sample size was determined by thematic saturation (19), whereby data collection ceased when no new themes or insights emerged from additional interviews. A total of twenty-two (22) participants were recruited and interviewed for this study.

### Data Collection

Data were collected between July and September 2024 using a semi-structured interview guide (20) (see supplementary file 1). This approach allowed for conversational flexibility while ensuring core areas relating to facilitators and barriers to dietary practices/behaviours. The interview guide was validated with three non-participating older adults to ensure clarity before its final application. Interviews were conducted in person at participants’ homes. While English was used for most interviews, seven participants preferred to be interviewed in Esan, a widely spoken local language. These sessions were audio-recorded on an encrypted voice recorder, and the researcher, acting as the translator, directly translated the audio into English during transcription, which was verified by an external person (E.J.I.) to ensure interpretation accuracy (21). The researcher maintained a detailed field journal to document contextual occurrences, non-verbal cues (body language), and personal reflections immediately following each interview.

#### Trustworthiness and Ethical Considerations

Ethical approval for the study was obtained from the research ethics committees of the School of Medicine and Population Health, University of Sheffield (Reference: 057009). Additional approval was received from Ambrose Alli University, Ekpoma, Nigeria (Reference number: HREC/20/24). Informed consent was obtained from all participants using signed consent forms, following a detailed explanation of the study with information sheet and opportunities to ask questions. Participants were explicitly informed of their right to withdraw at any time without providing justification or facing adverse effects; however, no participants withdrew during the study.

Study rigour was maintained through several measures: (i) Credibility was established via a detailed account of data collection and analysis and supported by the inclusion of direct quotations to allow readers to assess the link between evidence and interpretation. (ii) Fair dealing was practised by incorporating a diverse range of perspectives from the district (22). (iii) The analytical process actively searched for and investigated data components that contradicted emerging explanations to revise patterns. (iv) Monthly research team meetings provided a platform for peer debriefing and discussion on developing themes, further boosting the study’s credibility.

### Data Analysis

Interview recordings were transcribed by the lead author (O.D.I.) verbatim and checked by a second researcher (E.J.I.). The interviews that were not in English were translated into English O.D.I. and independently checked by E.J.I. The transcripts were thoroughly read to familiarise with the content before analysis. Data were analysed using a thematic analysis approach recommended by Braun and Clarke (23). This step involved a systematic six-phase process: (i) familiarisation with the data, (ii) generation of initial codes, (iii) searching for themes, (iv) reviewing themes, (v) defining and naming themes, and (vi) producing the final report. This method was chosen for its suitability in systematically identifying, organising, and interpreting patterns of meaning within complex social phenomena. The themes identified following the analysis were mapped across elements of the socio-ecological model (24). To preserve the integrity of participants’ voices, an inductive approach was also adopted, enabling the identification of supplemental themes that did not fit the predefined SEM categories, yielding a more nuanced and comprehensive final thematic map. NVivo 14 (QSR International) software was used to manage and organise the data throughout the analytical process (25).

## Results

### Participants’ characteristics

A total of twenty-two (*N* = 22) older adults aged 60 years and above participated in the study. The sample included equal numbers of men and women (*n* = 11 each). The majority were married (*n* = 16), while *n* = 6 were widowed. Eleven participants were retirees engaged in small-scale farming, 5 were petty traders, 4 were full-time farmers, and 2 were teachers. The ethnic composition was predominantly Esan (*n* = 14), with Bini (*n* = 3), Afemai (*n* = 3), and Igbo (*n* = 2). Data collection reached thematic saturation, confirming the adequacy of the sample. Participant characteristics are detailed in Table 1.

**Table 1.** Characteristics of study participants.

| **Participant Characteristic** | **Category** | **Frequency (*N* = 22)** |
| --- | --- | --- |
| **Sex** | Female | 11 |
|  | Male | 11 |
| **Marital Status** | Married | 16 |
|  | Widowed | 6 |
| **Occupation** | Retirees (with farming) | 11 |
|  | Trading (Petty Traders) | 5 |
|  | Farming (Full-time) | 4 |
|  | Teaching | 2 |
| **Religion** | Christianity | 16 |
|  | Traditional worship | 4 |
|  | Muslim | 2 |
| **Ethnic Group** | Esan | 14 |
|  | Bini | 3 |
|  | Afemai | 3 |
|  | Igbo | 2 |

### Facilitators and barriers analysis based on the socioecological model

Analysis of the interview transcripts identified 14 sub-themes, which were aggregated into four overarching themes mapped into the socioecological model: Individual, Environmental, Community, and Policy. The distribution of themes and sub-themes is summarised in Figure 1.

**Figure 1:**
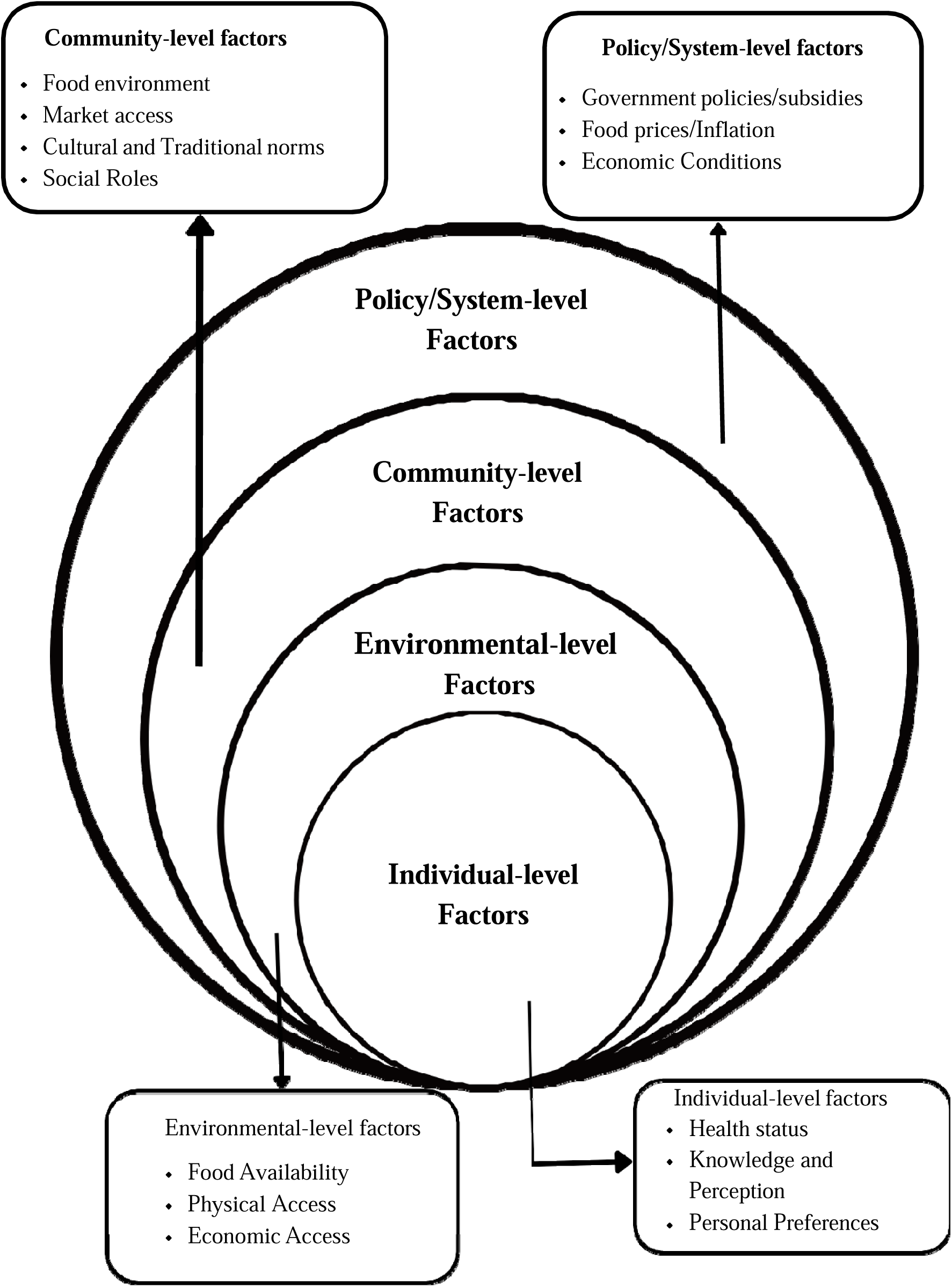
Factors influencing dietary choices/practices based on the Socioecological Model (SEM)

### Individual-level factors

#### Health status

Health-related factors presented both constraints and opportunities for change. Adapting to changes in diets facilitated dietary choices, for example, choices of food that would not lead to an increase in blood glucose for their diabetes condition, as participants recounted their adherence to medical advice in line with their health circumstances. Conversely, some participants reported that reduced physical strength, frailty, and mobility restrictions, which significantly limit their capacity to access food, shop, and prepare meals, were barriers to their dietary choices, leading them to choose options that require less preparation time or to buy what is available in nearby shops.

One of the most salient features of participants’ narratives was the progressive decline in physical capacity, especially in performing labour-intensive tasks such as food preparation, cooking, and food gathering. Even simple kitchen activities can become risky, with the potential for falls, burns, and other injuries. Such physical vulnerabilities reshape dietary practices, leading participants to rely on others, opt for less nutritious but easy-to-prepare foods, or skip meals altogether. Reduced physical capacity and frailty were direct barriers, making the logistical effort of meal preparation challenging. This was echoed by one female participant who said:

> *“My lack of strength makes it difficult to do all that is needed around the house each day… cooking and preparing meals have become a real challenge”* (P7, Female respondent)

The narratives also illustrate how chronic physical pain (especially joint, waist, and musculoskeletal discomfort) hampers mobility and food-related tasks. A female participant shared that:

> *“I’ve been having some joint pain lately that’s making it difficult to get around. The pain has been limiting my mobility and making it hard to move from one place to another… it makes getting access to my garden difficult”* (P6, Female respondent).

This kind of pain-related immobility severely reduces the ability to access markets, walk to farms, or even stand for extended periods while cooking. The loss of independence in these functions has deeper implications for psychosocial well-being, often leading to feelings of burden and influencing dietary choices/behaviours.

While following medical advice, adaptation to nutritional advice improved health outcomes, the required dietary restrictions often led to refraining from diets they are used to:

> *“I am managing high blood pressure, so I pay close attention to my diet and lifestyle to help stabilise my blood pressure levels”* (P19, Female respondent)

Within the socio-ecological framework, these individual-level health challenges intersect with broader structural and environmental factors to shape dietary practices and nutritional outcomes in later life. The findings reveal how age-related functional decline, chronic illness, physical pain, and fatigue directly and indirectly restrict autonomy over dietary choices, often resulting in reduced food intake, loss of dietary diversity, and increased vulnerability to malnutrition. Importantly, several participants also described the impact of diagnosed chronic illnesses on their dietary behaviour, reflecting a form of medically driven dietary adaptation. For instance, a female participant narrated:

> *“My dietary choices are significantly influenced by my health issues, particularly since I was diagnosed with diabetes… foods like garri and fufu… are now considered unsuitable for me due to their potential to exacerbate ulcer symptoms”* (P15, Female, respondent).

This statement reveals how health conditions introduce dietary restrictions that often conflict with local staple foods, thereby narrowing dietary choices. However, unguided dietary decisions may lead to nutrient imbalance, particularly when nutritional education or healthcare guidance is limited.

#### Knowledge and Perception

Participants demonstrated a robust, culturally grounded understanding of the health benefits of traditional, locally sourced foods. This deep-seated knowledge, stemming from lifelong experience, not only guides what they eat but also influences their dietary choices and behaviours, serving as a protective factor for maintaining health. To exemplify how the knowledge influences the dietary choices/behaviour, for example, diet choices and the high awareness of the fundamental role of nutrition, one participant stated:

> *“Nutrients play a crucial role in keeping the body and brain healthy, improving the functionality of our system”* (P1, Male respondent)

Consistent with this knowledge, the protective power of specific, well-known local foods was highlighted as an influence on the participants’ choice of diets:

> *“Adding vegetables is one of the best ways to boost the health profile of any meal. Greens like bitter leaf and pumpkin leaves are rich in vitamins, minerals, and fibre, and beans provide protein that helps build muscle and keep the body strong, while yams and cassava give energy”* (P11, Female respondent)

Participants consciously facilitated their dietary choices by maintaining a clear distinction between wholesome, natural, home-grown foods and modern processed options. This strong nutritional awareness led to the active avoidance of items such as noodles and refined sugars due to perceived chemical risks. The conscious effort to reject modern, artificial products in favour of traditional diets was articulated by one older woman:

> *“…As an elderly woman, I have learned to prioritise the nutritional value of my diet and steer clear of artificial and processed foods. I make a conscious effort to avoid items like noodles… Instead, I focus on traditional and wholesome foods”* (P16, Female respondent)

Despite general awareness, some participants held misconceptions rooted in their communities that influenced older people’s dietary choices. Such misconceptions are often associated with perceived personal intolerance or cultural axioms rather than scientific evidence. Participants reported that such misconceptions often led to unnecessary and self-imposed dietary restrictions that limited nutrient intake. The barrier created by perceived individual intolerance, which limits dietary variety, was exemplified by this comment:

> *“High-calorie foods, such as noodles and spaghetti, canned beef and sardines, are often characterised by their excessive calorie content relative to their nutritional value.”* (P22, Male respondent)

#### Personal Preferences

At the individual level, dietary choices were strongly influenced by personal taste and preference for familiar foods, which facilitated food selection. Conversely, food dislikes and aversions acted as barriers by constraining dietary variety. The primary role of immediate personal preference in driving food choices was stated plainly:

> *“My food choice depends on what I like to eat at a particular time”* (P12, Female respondent)

The narratives of the participants revealed that their dietary choices were primarily determined by what they liked, enjoyed, craved, and the taste of the food. The participants described their food intake based on their preferences. The taste of food determines their preference for one over the other, which in turn influences their decision. Some of the participants described their preferences based on the staples they enjoy most and what they would like to eat. Liking a diet was based on whether it tasted good or whether it was what they were craving.

> *“I base most of my food choices on two key factors: foods that I enjoy eating and foods that are healthy for my body”* (P3, Male respondent).

For some participants, their narratives about dietary preferences focused on what they enjoy and whether it is healthy for them. This means that if they do not like the taste or enjoy the food, they will discard it and avoid it. Some participants decided to have dietary aversions, especially when they felt it did not agree with their system. Others had food aversions, especially when the food was not satisfying or did not give them the energy they wanted.

One of the participants shared her experience, saying:

> *“I noticed that garri was not agreeing with me. Every time I ate it, I felt unwell, so I cut it out of my diet”* (P11, Female respondent).

### Environmental level factors

#### Food Availability

Participants consistently highlighted the availability of food as a critical environmental determinant influencing their dietary choices. This sub-theme reflects the dynamic interplay between local agricultural practices, seasonal variation, market accessibility, and broader socio-economic factors that shape food availability in rural communities. A recurring narrative among participants was that dietary consumption patterns among older adults were not solely determined by personal preferences but were largely constrained by what was available in their immediate environment. One respondent succinctly captured this sentiment, stating:

> *“We tend to consume what is readily available because, when options are at hand…”* (P18, Female respondent).

This view was echoed by others, who suggested that availability often superseded nutritional considerations or dietary diversity in shaping dietary choices and behaviours. Several participants reflected on the decline in local agricultural activity, which they associated with diminished variety and volume of food available in village markets. One female respondent recalled,

> *“In the past, there were numerous farmers bringing a wide variety of produce to the market. However, things have changed significantly. Nowadays, only a few people are engaged in farming, resulting in a much smaller supply of goods… we make do with what we see”* (P16, Female respondent).

This observation not only underscores reduced food diversity but also suggests a broader structural issue: the waning interest or capacity for subsistence farming among rural dwellers, which may be linked to ageing, youth urban migration, or economic disincentives. Perceptions of availability were closely intertwined with seasonality and economic constraints to shape dietary choices, as one male participant articulated,

> *“Generally, the food I eat is determined by what I can afford and what is readily available in my immediate surroundings. I rely on options that fit within my budget and can be easily sourced locally”* (P21, Male respondent).

Here, affordability emerges as both a filter through which availability is experienced and a limiting factor in their dietary choices and practices. Another participant echoed based on seasonality and said:

> *“Since it’s no longer yam harvest season, every time I visit the farm, I sip tea and eat bread, if any is available in the home, after brushing my teeth. Then I’ll make cocoa yams when I arrive at the farm”* (P9, Male respondent).

This anecdote reflects not only seasonal scarcity but also the adaptive strategies older adults employ to cope with fluctuations in food supply.

#### Physical access

Environmental insecurity, encompassing both climatic variability and sociopolitical instability, emerged as a critical constraint shaping older adults’ access to dietary practices in the rural context of Edo Central District. Participants consistently articulated how both natural environmental challenges and man-made threats had destabilised traditional agricultural routines, disrupted market supplies, and rendered subsistence farming increasingly precarious. A dominant concern was the impact of seasonal extremes, particularly the dry season, on crop availability. Respondents described the dry season as a period marked by scarcity and dietary compromise, in which dietary choices narrowed to what could survive under arid conditions. As one participant explained:

> *“There are times when a challenging season, such as the dry season, makes it impossible to obtain a lot of those food items. You then make the decision to harvest what you can….types of crops that can be cultivated in our area and the availability of certain ingredients also shape our dietary patterns”* (P1, Male respondent).

This statement reflects the adaptive strategies older adults employ in response to environmental stressors, often involving compromises in food variety and nutritional value. Here, food access is shown not to be merely a function of personal preference or economic means, but to be tightly bound to ecological limitations that regulate what can be grown, harvested, or even stored. However, the environmental challenges described by participants were not limited to climatic or agricultural conditions. A more severe and immediate threat was posed by insecurity due to violence and conflict, particularly involving the Fulani herdsmen insurgency. For several respondents, this sociopolitical instability had led to a complete cessation of farming, leading to food shortages and limited dietary options. One female participant described the gravity of this threat:

> *“Due to the security challenges posed by the Fulani herdsmen insurgency, I was forced to cease my farming activities. The ongoing conflict and instability in the region have made it increasingly dangerous and impractical to continue working on my farm. The threats to personal safety and the disruption of agricultural infrastructure have created an environment where farming has become too risky and uncertain”* (P15, Female respondent).

This testimonial shows how personal safety has become an overriding concern, often outweighing livelihood needs. What was once a stable practice (subsistence farming) has now become a high-risk endeavour due to physical threats, destruction of farmland, and psychological trauma. This generalised fear among community members has far-reaching implications for food security. As fewer farmers are willing or able to cultivate land, the agricultural supply chain contracts, reducing the volume and diversity of food supply available in local markets. As one woman explained:

> *“Sourcing food from the farm has become challenging. Many farmers are no longer bringing their produce to market, resulting in a limited supply”* (P18, Female respondent).

These disruptions to market supply, due to security concerns, compound existing vulnerabilities linked to poverty, ageing, and climate change. In this context, dietary choices/practices become both environmental and political problems, ones that cannot be resolved solely through individual coping strategies. These insights reflect the interconnectedness of ecological, economic, and security dimensions of access to food and influence on dietary choices/practices. The combined effects of climate variability, seasonal food shortages, and regional insecurity create a fragile, uncertain food environment for older adults, many of whom lack the resilience (physical, financial, or social) to navigate these shifting risks.

#### Economic Access

Affordability, or the ability to economically access a sufficient and diverse range of food, emerged as one of the most prominent environmental-level influences on the dietary choices/practices of older adults. Participants repeatedly referenced the limitations imposed by financial insecurity, often describing constrained dietary choices and compromised nutritional quality as direct consequences of economic hardship. These accounts revealed that affordability was not merely a matter of food prices but was tightly interwoven with broader structural vulnerabilities, including widowhood, inflation, inconsistent farm income, and inadequate social support systems. A recurring narrative was the notion of budget-driven consumption, in which participants’ dietary choices were not necessarily what they preferred but what they could afford. Some participants illustrated this point clearly:

> *“Generally, the food I eat is determined by what I can afford and what is readily available in my immediate surroundings. I rely on options that fit within my budget and can be easily sourced locally”* (P21, Male respondent).

This statement highlights the economic rationalism that governs dietary choices among older adults, in which affordability constrains autonomy and choice. It also reveals a dual limitation, such as financial (budget) and spatial (locality), that jointly mediate access to food. Several participants framed affordability in clear, basic terms of necessity. Rather than discussing dietary preferences, they spoke of price sensitivity, tactical buying, and a constant effort to “*make do*”. That indicated cost eclipses nutritional consideration in most cases. For many, the ability to “*eat at all”* was prioritised above the ability to “*eat well”*. This reflects the logic of food- and diet-related coping strategies, in which affordability serves as the core axis around which all dietary decisions pivot. The experiences of female participants, particularly widows, shed light on how affordability intersects with gendered economic disadvantage. One widow reported,

> *“Life has been difficult since losing my husband. As a widow, I find myself struggling financially on a daily basis… so I’ve had to make some adjustments to my diet”* (P10, Female respondent).

Her narrative not only foregrounds her uncertain economic situation but also indicates the decline of household income and social protection following bereavement. In contexts where older women are often economically dependent on their spouses and face restricted access to formal employment or pensions, widowhood becomes a critical turning point for dietary choices/practices, leading to nutritional vulnerability. Such adjustments may involve reductions in portion sizes, meal frequency, or food quality, with implications for the nutritional adequacy of older adults’ diets. Other respondents echoed the theme of constrained dietary diversity. One female participant noted:

> *“Limited financial resources restrict my ability to buy the foods I desire, making it difficult to access and enjoy a diverse range of options”* (P18, Female respondent).

This reflects a conscious awareness of nutritional aspiration, that is, participants were not unaware of what constituted a diverse and healthy diet, but were unable to achieve it due to persistent financial shortfalls. The quote also reflects the psychological burden of unmet dietary expectations, a subtle but significant form of food-related stress. This shows that participants knew what constituted a “good” or “healthy” diet but identified affordability as the principal barrier to achieving it.

### Community Level Factors

#### Food environment

Food sourcing among older adults emerged as a central determinant of dietary choices/practices, with participants overwhelmingly identifying subsistence farming as their primary means of acquiring food/diets. This sub-theme highlights the centrality of self-grown produce in respondents’ diets and the secondary, supplementary role of local markets. A dominant narrative among participants was that most of the food they consumed was grown directly, either on small personal farms or in home gardens. As one male respondent stated,

> *“Typically, I can obtain most of my food directly from the farm. However, in rare instances when the farm’s supplies are depleted, I must resort to purchasing items from the market”* (P3, Male respondent).

This response highlights both the self-reliance fostered by farming practices and the adaptive use of markets as a secondary food source when harvests are insufficient as factors influencing dietary choices/practices within the participant’s domain. Similarly, another participant elaborated,

> *“Most of my food comes from my own farm. I only buy things that I can’t grow or raise myself, like rice and beans. My little farm gives me most of what I need to survive”* (P4, Male respondent).

This reflects a strategic balance between agricultural production and selective market dependence, where non-locally grown or non-staple foods are purchased when necessary. Several respondents also described a hybrid model in which they utilised both farm produce and market goods, often dictated by seasonality, crop yields, and access. One participant stated,

> *“I regularly make trips to the bustling open-air market located in the centre of town. In addition to supplementing my meals at the market, I am also fortunate to have my own modest plot of land where I cultivate crops”* (P5, Male respondent).

This dual sourcing approach enabled greater dietary variety and a buffer against the unpredictability of subsistence farming. The overwhelming reliance on farm produce was reiterated throughout the Participants. These assertions reflect the deeply embedded agrarian lifestyle in rural areas, where farming is not only a livelihood but also the principal means of food security. These not only underscore the cultural and sensory value placed on farm-grown food but also point to a nutritional advantage: fresh, unprocessed, and self-grown foods are regularly consumed. While market food was not the dominant source, it still played an important role, particularly in filling dietary gaps or supplementing during off-seasons. Thus, the frequency and productivity of farming directly influenced market dependence. Seasonality also shaped sourcing practices. A participant observed,

> *“Our food choices typically depend on what we grow on the farm. We also occasionally purchase items from the local market. We have access to seasonal produce, and we tend to consume more of those foods during their respective growing seasons”* (P3, Male respondent).

This reflects a broader ecological rhythm in which agricultural cycles determine the types and quantities of diets consumed at different times of the year. This suggests that while farming provides a degree of autonomy, economic realities still influence what is purchased and consumed beyond self-grown staples. While subsistence farming was the primary means of food procurement for many participants, there was a clear and consistent pattern of market dependency, particularly for essential staples not cultivated locally.

#### Market Access

Community markets occupy a central and enduring position within the lived dietary choices/practices of older adults. These markets function not merely as transactional spaces but as deeply embedded socio-cultural institutions that structure routines, relationships, and access to dietary resources. One of the most salient dimensions of this sub-theme was the predictability and regularity of market occurrences. Participants reported aligning their household dietary management strategies with cyclical market schedules, with most markets operating on a 4-day cycle. This rhythm, deeply woven into local temporal structures, facilitated both planning and provisioning, especially in households where subsistence agriculture alone could not sustain a diverse or adequate diet. As one respondent noted:

> *“Our community has established specific market days. In our area, there is a designated market day every four days. On these scheduled days, I make a point of visiting the market to buy the items I need. This routine helps me stay stocked up on essentials and manage my household needs effectively”* (P18, Female respondent).

This suggests that community markets are not perceived as irregular or discretionary; instead, they are integral to the temporal and material organisation of food access and influence on dietary choices/practices in rural households. The markets serve as planned interventions against scarcity, enabling households (especially those with limited garden yields) to replenish their supplies in a consistent and manageable way. Beyond logistical considerations, the markets were also described as socially embedded and relationally meaningful spaces. Long-term engagement with specific vendors led to the cultivation of trust, social capital, and occasionally economic advantages in dietary choices. As some participants explained:

> *“I’ve been going to the same market for years, and at this point I know many of the vendors quite well. They are always happy to give me a good deal”* (P11, Female respondent).

This familiarity was not merely incidental but formed part of a strategic relational economy, in which enduring interpersonal ties could influence the pricing, quality, and reliability of food access and dietary choices. The market’s social infrastructure thus facilitated dietary choices for older adults, many of whom faced mobility, income, or declining agricultural output constraints. Further, the markets were perceived as responsive to seasonal and supply dynamics, with participants adjusting their market behaviour in line with crop cycles and product availability. For example, one participant highlighted the value of staying informed about the weekly fluctuations in fresh food supply:

> *“Every market day, I like to go and get the food I need. The local open-air market is held weekly, and it’s always good to see what fresh foods are available that week”* (P4, Male respondent).

These routine-based food access strategies reflect a tightly calibrated interplay between personal agency and structural availability, whereby older adults actively monitor and adapt to the shifting contours of the rural food landscape.

#### Culture and Traditional norms

Culture emerged as a deeply embedded and enduring determinant of dietary choices/practices among the older adults, shaping not only the kinds of diets consumed but also the meanings ascribed to eating and meal preparation. Participants conveyed a strong attachment to local food traditions, often viewing their dietary choices/practices as inseparable from their cultural heritage. In this context, traditional diets were not merely sustenance but a powerful expression of identity, continuity, and belonging. Several participants emphasised the centrality of culture in shaping their dietary choices/behaviours, how food was prepared, and the values associated with eating. As one male respondent observed:

> *“The foods that have been locally eaten here for generations influence the choices I make about what to cook and eat. The culinary traditions passed down from earlier times still shape my relationship with food today…the food that is mostly produced in this region of the world is so addictive”* (P9, Male respondent).

Here, the term “addiction” should be interpreted not as a clinical dependency, but as a colloquial expression of deep-seated cultural preference and habitual familiarity with local staples. This illustrates how cultural embeddedness can promote a form of dietary resistance, where traditional foods are repeatedly chosen not simply for taste or affordability but because they resonate with collective memory and identity. Similarly, another respondent described these locally produced foods as “so addictive”, reinforcing the idea that dietary habits in the community are more than routine; they are internalised and emotionally resonant. This pattern reflects dietary socialisation from early childhood, in which individuals become conditioned to specific tastes, textures, and preparation styles that continue to influence their dietary choices across the lifespan. The intergenerational transmission of food preferences was another recurring theme. One male participant explained:

> *“My cultural background has had a profound influence on me, especially when it comes to food. The meals I was raised with as a child have stayed with me into adulthood and really shape the kinds of foods I enjoy”* (P5, Male respondent).

These quotes underscore the longitudinal continuity of dietary practices, in which dietary norms established in early life persist into older age, even amid changing socio-economic and environmental conditions. This continuity is particularly salient in rural communities where modernisation and globalisation may be less intrusive, and traditional food systems remain robust. Importantly, participants associated these traditional diets with healthfulness and sustainability, echoing sentiments that local diets were not only familiar but also nutritionally beneficial. One female respondent highlighted the nutritional value of indigenous staples:

> *“Preparing homemade meals using fresh, local ingredients is so important for your health and well-being”* (P12, Female respondent).

Although the classification of food by macronutrient content may appear simplistic, it reflects a folk nutritional knowledge that often aligns with biomedical understanding, particularly in contexts where access to formal dietary counselling is limited. In addition to preferences and perceptions, participants spoke of food preparation practices as integral to their cultural worldview. This highlights a holistic understanding of diets that incorporates both the material (nutrients, freshness) and the processual (cooking, preparation, selection) dimensions. Food/diets were seen not only as products but also as processes intimately tied to the local environment and traditional knowledge systems. This speaks to the historicity of dieting, the way in which current eating practices are rooted in a temporal continuum, shaped by the recipes, practices, and ingredients handed down through generations.

#### Social roles

This subtheme describes household decisions regarding meal consumption and preparation. It shows that household dynamics influence these choices. Based on participants’ stories and descriptions, it was evident that families reach a collective agreement on the daily diet. Although the decision about what to eat is collective, the data indicated that women mainly handle meal preparations. This revealed that the norms in the region hold that women prepare meals, which influences diet choices. Participants also revealed that the person preparing the meals sometimes makes decisions without consulting other family members. For the individual participants who live alone, they made their decisions as they would not need to seek consent from anyone.

> *“My wife, as the primary cook in our household, is responsible for deciding what food we eat. Since she prepares all the meals consumed in our home, she has the main decision-making role when it comes to our diet”* (P3, Male respondent).

On the other hand, participants also explained that their dietary preferences were not taken into consideration, even though they may have had a choice in what to eat. They explained that since they are not the ones who plan the meal, the person who plans and prepares it makes the overall decision. One of the participants’ explanations is captured below:

> *“I have to admit that my wife usually makes the final decision about what to eat. She usually plans our meals, even if I might have some preferences”* (P4, Male respondent).

### Policy/System-level factors

#### Government policies (Subsidies)

A dominant and recurring concern among participants was the perceived lack of meaningful government support to ensure food security and enhance agricultural productivity. Across participants, older adults expressed deep dissatisfaction with the government’s failure to provide structural, financial, or policy-driven assistance, particularly in rural and agrarian communities. The lack of direct intervention in agriculture (such as subsidies, grants, or input support) was a major factor influencing participants’ dietary choices. Respondents repeatedly stated that, despite the centrality of farming to their dietary practices, they had never received any assistance from the state. One male participant underscored the lifetime absence of state support, stating:

> *“All my life, from the day I was born until now, I have never benefited from any kind of government assistance. I have never received a subsidy, grant, or any other financial support”* (P9, Male respondent).

This statement powerfully illustrates the systemic exclusion of rural farmers from policy benefits, despite the state’s public declarations of its commitment to agricultural development. Similarly, another respondent noted:

> *“They do not provide any assistance and do not lower the cost of food supplies; therefore, they have no influence. They are not our farm’s financiers”* (P1, Male respondent).

This reflects a strong perception that the state not only fails to support local food producers but also remains detached from the structural realities of rural agriculture. The lack of financial incentives or subsidies for agricultural inputs hinders efforts to improve production diversity or scale, thereby leading to insufficient variety from which to choose. The inadequacy of institutional regulation, particularly in controlling food prices and ensuring availability, was another significant complaint. Multiple participants linked the soaring cost of living to the absence of government price control mechanisms, with many voicing concerns that the state had abandoned its responsibility to protect low-income households from inflationary shocks.

Despite the state’s promises to address these economic challenges through policy, participants emphasised that the interventions were either symbolic or poorly implemented. As one female respondent lamented:

> *“The government is not contributing to improving our nutritional needs. They fail to implement effective policies that could benefit us and instead make things more difficult. Rather than introducing measures that could support us, they impose additional burdens and take away resources that could otherwise be used for our benefit”* (P16, Female respondent).

This perceived failure of policy delivery mechanisms reinforces a broader narrative of disillusionment and policy alienation. Some participants went further, accusing the government of actively exacerbating hardship through counterproductive policies. In a few cases, respondents described a complete absence of government presence in their communities. This was framed not just as policy neglect but as a total abdication of responsibility, especially toward farmers. Taken together, these narratives reveal a profound disconnect between policy intentions and community realities. While the government may espouse food security and agricultural revitalisation in official rhetoric, the absence of tangible, equitable, and targeted support mechanisms fosters widespread distrust, alienation, and cynicism among the rural older adults.

#### Food Prices/Inflation

A factor that emerged within the policy domain is the pervasive impact of food price fluctuations, which participants cited as one of the most destabilising forces affecting food access and dietary choices. Respondents across genders and regions emphasised that rising, unpredictable prices have significantly constrained their purchasing power, making once-affordable staples inaccessible and eroding household food security. Several participants attributed these fluctuations directly to poor government regulation and policy failure. One respondent articulated this link clearly, saying that rising food prices were perceived as resulting from both economic instability and weak regulatory oversight:

> *“There is a lack of effective regulation and management when it comes to food prices and availability. While the government may be making efforts to address these issues, many people feel that these efforts fall short of what is needed”* (P21, Male respondent).

The failure to control inflationary pressures and support agricultural supply chains has led to dramatic increases in staple food prices. For instance, rice and garri, once widely consumed and relatively inexpensive, have become luxury items for many:

> *“The cost of a bag of rice has increased so much that what used to buy a full bag now only covers half. Similarly, garri, once considered an affordable staple for the average person, is now priced around LJ80,000”* (P13, Male respondent).

This sharp escalation in food prices is not just an economic inconvenience but a critical threat to dietary choices and food access, particularly for low-income and vulnerable populations. Respondents reported having to strategically reduce the quantity and variety of foods they purchase in response to these increases. The strain extends beyond altered purchasing habits to overall food insufficiency. These economic shocks have direct nutritional consequences, particularly for older adults who may already have limited incomes and specific dietary needs. For instance:

> *“Food prices are increasingly becoming unaffordable. For instance, I should be eating beans as part of my diet, but the cost is too high for me to manage. The continual rise in food prices makes it challenging to maintain a balanced diet, as essential foods become out of reach, impacting my ability to meet my nutritional needs”* (P20, Female respondent).

These reflections highlight how unregulated food markets and erratic pricing patterns disproportionately affect nutritionally vulnerable groups. The volatility in food prices not only limits access but also undermines efforts to plan for healthy, sustainable diets. Indeed, price volatility was not only experienced as a chronic issue but as a daily source of uncertainty and anxiety. The dynamic nature of local pricing was captured vividly in one participant’s comment. This quote illustrates the instability of the food economy in many communities, where residents must constantly adapt their diets to reflect fluctuating market realities. Such unpredictability erodes confidence in food systems and contributes to long-term food insecurity, particularly among those already facing socioeconomic disadvantage.

#### Economic conditions

Financial instability emerged as a pervasive barrier to dietary choices and diverse food access among older adults. Across multiple narratives, respondents described how chronic financial hardship, exacerbated by inflation, economic stagnation, and limited income-generating opportunities, shaped not only the quantity and quality of dietary choices/behaviours but also their emotional and psychological relationship with food acquisition. One of the most comprehensive articulations of this issue came from a male respondent who emphasised the dynamic nature of financial capacity in shaping dietary patterns:

> *“Certainly, money is a crucial factor that greatly influences my food choices. My financial capacity determines the type and quality of food I can afford. When funds are limited, I have to prioritise more affordable options, which can impact the variety and nutritional value of my diet. Conversely, when my financial situation allows, I have more flexibility to choose healthier and more diverse food options”* (P1, Male respondent).

This statement encapsulates the duality of dietary choices/behaviours/practices, oscillating between nutritional adequacy and deprivation, depending on one’s financial context. However, for most participants, the reality leaned heavily toward persistent economic shortfall. Several women described a growing sense of powerlessness amid rising costs and shrinking purchasing power. Here, inflation is directly linked to dietary insufficiency, as even consistent income fails to match the escalating costs of basic food items. The inability to meet prescribed nutritional needs was another recurring theme, especially among participants managing chronic health conditions. For example:

> *“I don’t have the financial resources I need. Whenever I receive a prescription for the foods I should eat, I often feel distressed because I can’t afford them”* (P20, Female respondent).

This quote reveals how economic deprivation directly compromises medical compliance, placing older adults at heightened health risk. Financial hardship thus intersects with healthcare inequities, reinforcing cycles of food insecurity and poor health outcomes. Daily dietary choices were described as acts of budgetary negotiation, where desires were subordinated to available funds. Some participants linked their financial precarity to insufficient or unstable income, often tied to a lack of employment or limited agricultural sales. One respondent noted:

> *“Unfortunately, I do not currently have enough funds to purchase the food I wish to consume. If I do not sell some of my agricultural produce, I will lack the financial means to procure provisions from the local marketplace”* (P9, Male respondent).

For many, the financial situation had deteriorated so significantly that they began buying food in smaller quantities, reflecting constrained liquidity. This shrinking consumption pattern has implications not only for food security but for mental well-being, particularly when past patterns of food abundance contrast sharply with current austerity. Finally, participants like this woman summarised the overall economic pressure by expressing a longing for financial sufficiency. These narratives underscore that for older adults, dietary choices are not primarily shaped by preferences, cultural traditions, or even availability, but by money. The data reveal that financial instability does not simply limit access to food; it redefines the entire process of acquiring, preparing, and even imagining meals.

## Discussion

This study explored the facilitators and barriers influencing dietary choices/practices among older adults living in rural communities in Edo Central, Nigeria. Using a qualitative, exploratory approach guided by the socio-ecological model, semi-structured interviews were conducted with 22 older adults aged 60 years and above. The interviews provided detailed, contextualised insights into the complex factors shaping dietary behaviour in later life. The data were analysed thematically, following Braun and Clarke’s method (23), identifying interconnected factors across individual, environmental, community, policy/system levels. The key findings are discussed below.

### Individual factors

Health status emerged as a key individual-level determinant shaping dietary choices among older adults. It highlights how age-related physiological decline interacts with broader structural constraints to influence dietary behaviours and food security in later life. As functional capacity declines with age, routine tasks such as food procurement, preparation, and cooking become increasingly demanding, thereby limiting older adults’ ability to maintain preferred dietary patterns (17). These physical limitations are not experienced uniformly. In many contexts, older women continue to bear primary responsibility for domestic labour, including food preparation, which may exacerbate physical strain and reduce their autonomy in making food-related decisions (26). This gendered dimension of ageing underscores how individual health status is closely intertwined with social roles and expectations.

In response to declining physical capacity and emerging health conditions, many participants described actively modifying their dietary practices. These adjustments were frequently framed as deliberate strategies to manage chronic diseases such as hypertension and diabetes. Participants reported adhering closely to clinical advice, adopting dietary practices aimed at stabilising blood pressure or controlling glycaemic levels (27). Such practices included reducing salt and sugar intake, modifying portion sizes, and prioritising foods perceived to support disease management. These findings suggest that dietary decision-making in older age is increasingly shaped by the management of chronic conditions, reflecting a shift from food choice based primarily on preference or tradition towards medically informed dietary regulation.

Importantly, this proactive engagement with clinical guidance indicates that older adults are not passive recipients of health advice but active agents who adapt their dietary behaviours in response to changing health needs. However, the ability to implement such dietary adjustments may still be mediated by broader structural factors, including access to appropriate foods, financial resources, and social support (28). This underscores the need to consider health status not only as an individual determinant but also as a factor embedded within wider socio-ecological contexts that shape food access and dietary practices among older populations.

Beyond health, awareness of nutritional benefits and personal food preferences were consistently highlighted as important influences on dietary choices among older adults. Participants demonstrated considerable knowledge of the perceived health benefits of different foods, often linking specific dietary practices to improved strength, satiety, and general well-being. Many showed strong familiarity with locally available nutrient-dense foods and were able to associate these foods with food groups and health outcomes. This awareness appeared to encourage a preference for foods perceived as “healthy,” typically described as locally grown, natural, or minimally processed, while refined sugars and highly processed foods were consciously avoided. Traditional staples such as *fufu* were frequently valued for their perceived ability to provide sustained energy and promote longevity, reflecting the continued cultural importance of traditional diets in supporting health in later life. These findings are consistent with the Food Choice Process Model (28) which emphasises how personal food systems, comprising accumulated knowledge, cultural meanings, and lived experiences, influence everyday food decisions.

However, dietary choices were not always guided solely by evidence-based nutritional knowledge. Participants’ perceptions of food healthfulness were sometimes shaped by culturally embedded beliefs and personal experiences, including a strong preference for homegrown foods and the avoidance of certain foods following negative digestive experiences. Within the Socio-Ecological Model, these patterns illustrate how individual-level factors such as knowledge, beliefs, and sensory preferences interact with cultural norms and local food environments to shape dietary behaviours (24, 29, 30). Taste and sensory satisfaction remained important drivers of food choice, highlighting that dietary decisions in later life reflect a complex interplay between nutritional awareness, cultural interpretations of food, and embodied experiences of health and wellbeing (30). These findings underscore the need for culturally sensitive nutrition interventions that build on existing knowledge while addressing misconceptions that may unintentionally limit dietary diversity among older adults.

### Environmental factors

In this rural context, food availability was a primary determinant of dietary choices/practices, shaped by local production cycles and geographic accessibility, a finding that aligns with previous studies in Nigeria and Sierra Leon (31, 32). In Sierra Leon, seasonal fluctuations largely dictated dietary adaptations during “lean times” or off-harvest periods. Households often replaced traditional staples with less nutritious alternatives, a shift that markedly reduces dietary diversity (32). All these findings suggests that in regions where local production is highly seasonal, older adults must constantly recalibrate consumption in response to the fluctuating availability of nutrient-dense foods (33). As one participant noted, *“options are mostly restricted to what is readily available,”* highlighting how systemic limitations in market infrastructure and agricultural output supersede individual agency (32).

Participants noted that physical access to food was increasingly undermined by a dual threat of climatic variability and sociopolitical instability. Beyond this, the predictable challenges of the dry season, further fuelled by insurgency of herdsmen was highlighted as a critical layer of food insecurity (34). Conflict-driven farm abandonment has halted subsistence production, the region’s dominant food source, thereby reducing choices and escalating the risk of acute malnutrition among older populations who maintain deep cultural and economic ties to agrarian lifestyles (35).

Economic access was another determinant, with affordability influencing dietary choices/practices, forcing a prioritisation of caloric quantity over nutritional quality. Participants consistently described a survivalist approach to consumption, characterised by the mandate of “eating at all” rather than “eating well” (36). This financial precarity is often exacerbated by intersecting factors, including widowhood, the lack of robust social protections, and the volatility of agricultural income (37). To manage these constraints, older adults employ various coping mechanisms, including reducing portion sizes or liquidating surplus produce to fund essential market purchases. These strategies illustrate that the affordability of diets is a multi-dimensional challenge involving both financial and spatial constraints (38). Furthermore, these economic barriers are gendered; older women, particularly widows, face heightened vulnerability following the loss of spousal income, which frequently results in a direct decline in nutritional adequacy (37).

### Community factors

Community-level factors, including the local food environment, market access, and prevailing cultural norms, emerged as important influences on older adults’ dietary choices. In many rural African settings, subsistence farming continues to represent a foundational pillar of household food security, particularly for older populations whose livelihoods remain closely tied to agrarian practices. Our findings indicate that home gardening and small-scale farming provide an important source of dietary diversity while reinforcing long-standing cultural practices linked to land, food production, and self-reliance (39, 40). Similar observations have been reported in other rural African contexts, where subsistence agriculture functions not only as a food provisioning strategy but also as a means of sustaining traditional diets and buffering households against fluctuations in food prices and supply (40). However, the inherent unpredictability of rain-fed agriculture means that farming alone rarely satisfies all household dietary needs. Consequently, households often rely on a hybrid sourcing strategy in which self-produced staples form the dietary foundation while local markets serve as complementary sources for non-staple foods and seasonal shortages (41). This dual strategy reflects a pragmatic adaptation to environmental and economic uncertainties and has been documented across several African food systems where smallholder production and informal markets operate as interdependent components of local food environments (41, 42).

Local markets play a particularly important role in shaping dietary practices among older adults. Beyond their function as sites of food exchange, rural markets serve as important socio-cultural institutions that structure food access, dietary planning, and social interaction within communities. Regular market cycles enable households to plan food purchases and diversify diets, thereby expanding access to foods that may not be locally produced (43). Moreover, the social relationships that develop between vendors and consumers often function as informal safety nets, particularly for older individuals with limited mobility or financial resources. In many African settings, these interpersonal relationships facilitate flexible pricing arrangements, informal credit, and preferential access to fresh produce, thereby helping to sustain food access among vulnerable groups (44). However, these traditional market systems are increasingly under pressure due to broader structural changes in rural economies. The outmigration of younger generations to urban centres, declining agricultural productivity, and shifting food supply chains have been associated with reduced diversity of foods available in local markets (45). As a result, scholars have increasingly emphasised the importance of strengthening rural food environments, through improved infrastructure, support for smallholder agriculture, and revitalisation of local markets, to enhance dietary diversity and nutritional outcomes among ageing populations (41).

Dietary choices were also strongly shaped by cultural norms and social meanings attached to food. In many African contexts, food consumption extends beyond meeting physiological needs and is closely tied to cultural identity, social belonging, and intergenerational continuity (46). Participants’ descriptions of certain traditional foods as “addictive” reflected not a literal dependence but rather a deep emotional and symbolic attachment rooted in cultural familiarity and shared culinary traditions. These findings align with existing literature suggesting that traditional diets often function as markers of cultural identity and collective memory, reinforcing continuity between generations and maintaining social cohesion within communities (46). Similarly, perceptions of food quality and healthfulness were strongly influenced by food provenance. A pronounced preference for self-produced or farm-grown foods reflects widespread cultural beliefs that locally produced foods are more natural, pure, and trustworthy than commercially sourced alternatives (47). Such perceptions may partly stem from concerns about food safety and the industrialisation of food systems, but they also reflect longstanding cultural values associated with land, farming, and traditional food preparation practices.

Importantly, these findings highlight how community knowledge systems and cultural narratives shape dietary practices in ways that may both support and challenge evidence-based nutritional guidance. While traditional diets in many African settings are often rich in whole foods and minimally processed ingredients, reliance on culturally embedded knowledge may also lead to the persistence of beliefs that are not always aligned with current nutritional recommendations. As such, nutrition interventions aimed at improving dietary outcomes among older adults must move beyond purely biomedical messaging and instead engage with the cultural meanings, social relationships, and local food environments that shape everyday food choices. Culturally sensitive approaches that recognise the role of traditional food systems and community knowledge are therefore essential for promoting sustainable dietary improvements among older populations in rural African contexts (47, 48).

### Policy/System factors

Participants identified a multifaceted array of structural barriers to dietary adequacy, ranging from macroeconomic volatility and inflation to perceived inadequacies in state interventions. A primary theme emerged regarding the decoupling of state rhetoric from agricultural reality; while the government publicly asserts a commitment to agricultural development, respondents consistently reported a total absence of direct state assistance (49). This systemic exclusion undermines the sustainability of rural livelihoods. Scholarly literature suggests that when subsidies are effectively channelled into infrastructure, e.g. electricity or credit facilities, it signals a protective state intervention aimed at ensuring national food security (50). However, the lived experiences of these respondents, characterised by assertions that they are “not our farm’s financiers,” underscore a significant gap in policy implementation. While certain regional initiatives, such as agricultural insurance, aim to mitigate production risks (51), the lack of direct fiscal support limits farmers’ ability to diversify crops or scale operations, leading to fewer dietary options. This discrepancy suggests that resource allocation may be determined by extraneous political factors rather than the immediate caloric or financial needs of food producers (50, 52). The resulting financial constraints force a stagnation in productivity, with profound implications for long-term rural development.

The current study data further highlight an institutional failure to regulate food prices as a barrier to dietary choices. Participants linked erratic pricing patterns to weak regulatory oversight and market forces that have rendered staples, such as rice and garri, increasingly inaccessible to the older adult demographic (53). Despite official stabilisation mandates, these mechanisms remain largely ineffective for low-income households. This misalignment between policy goals and outcomes points to broader institutional fragility, in which regulatory frameworks fail to buffer the effects of global trade dynamics and domestic instability (54). Empirical evidence indicates that effective price control necessitates cross-sectoral policy coherence (55). In the current study, participants specifically noted that *“*governmental policies have put pressure on farmers,*“* inadvertently triggering market spikes (56). This corroborates existing research: in the absence of targeted state-level intervention, food markets become highly volatile, precipitously eroding the purchasing power of vulnerable consumers (57).

Participants’ economic conditions were another determining factor influencing dietary choices/behaviours. Rising costs necessitate a shift in household procurement strategies, often leading to a “strategic reduction” in food expenditure (58). This coping mechanism prioritises caloric volume over nutritional density, directly compromising diet quality. In the Nigerian context, such unpredictable pricing cycles push households into a state of “sporadic scarcity,” compounding wider economic instability (58, 59). From a theoretical perspective, limited monetary resources serve as the primary determinant of food choice. Respondents consistently prioritised affordability over nutritional adequacy, a behaviour that aligns with extended applications of the Theory of Planned Behaviour (TPB) (60).

### Strengths and Limitations

A primary strength of this study lies in its use of an in-depth, qualitative approach, which provided rich, context-specific insights into the complex, interconnected determinants of dietary choices within a poorly researched population in rural Nigeria. The use of the socioecological model ensured a holistic analysis, moving beyond individual behaviours to expose the systemic policy, environmental and community barriers that restrict dietary adequacy. The inclusion of the local language, Esan, in the interview process enhanced the credibility of the data.

However, the findings are subject to limitations. As a qualitative study, participants’ responses may have been influenced by social factors, particularly given the nature of the questions. However, they are highly relevant to similar rural, agrarian communities in the region. Furthermore, the data rely on self-reported perceptions of dietary choices, consumption and health, which may be subject to social desirability bias or recall error.

## Conclusion

The dietary choices and practices of older adults in rural Edo Central are determined by a complex interplay of internal resilience and external structural fragility. While their deep cultural knowledge facilitates healthy intent, this is consistently overridden by macro-level barriers, including pervasive financial instability, food price volatility, and the acute threats posed by insecurity and environmental change. The knowledge of nutrition and the perception of healthy eating among older adults in the rural context in this study revealed that they would maintain healthy eating practices when they had abundant access to healthy foods. Addressing food security and optimal nutrition for this vulnerable population requires integrated policy action that strengthens the local food system, provides economic protection, and offers culturally resonant health education. Continued research should focus on quantitatively measuring the impact of these policy failures on nutritional status and health outcomes.

## Declarations

## Human Ethics and Consent to Participate

Ethical approval was obtained from the University of Sheffield (Reference number: 057009) and Ambrose Alli University (Reference number: HREC/20/24).

## Consent to Publish declaration

Not applicable

## Data Availability Statement

Data from this study are available and can be obtained from the corresponding author (Onosolesena Idiakheua- and the following co-authors: Robert Akparibo-, Elizabeth Williams-, Viren Ranawana-, upon a justifiable request.

## Acknowledgement

The authors express their gratitude to the study’s participants for their time and effort. The authors are grateful to the research assistants for their time and dedication during data collection for this study, and to the Tertiary Education Trust Fund (TETFund) for its support. The authors appreciate the University of Sheffield Open Access for funding the publication costs of this study.

## Authors Contributions

Onosolesena Dennis Idiakheua (O.D.I.) conceptualised the study with advice from R.A., V.R., and E.A.W. O.D.I. wrote the first draft with significant input from R.A., V.R., and E.A.W. AA supported fieldwork and data analysis. E.J.I. verified the translation and transcription. O.D.I., E.A.W., V.R., and R.A. reviewed the manuscript. All authors proofread and approved the final version. E.A.W., V.R., and R.A. supervised the study.

## Supplementary file

### QUALITATIVE STUDY

#### In-depth Interview Guide (IDI)

##### Introduction and Warm up (2 mins)

***Moderator Note: Moderator to check participants are okay to participate and informed consent is obtained (signature/ thumb print)***

⍰ The moderator introduces him/herself and explains the purpose of the study.
⍰ The moderator explains that the discussion is open, not an exam and there is no right or wrong answer. Explain that the information given by the participant is confidential.
⍰ Encourage the respondent to give honest opinions.
⍰ Explain the use of the recorder.
⍰ The target group that the interviewee falls under must be documented.
⍰ Start tape recording if consent is granted: (Moderator to switch recorder on)

**I will ask you questions about your food intake, nutrient intake, nutrition practices and what influences the choices you make.**

1. Can you tell me about your diet/foods that you eat? What do you think influence your food choices? Is it personal preferences? Cultural background? Taste? Financial status?

i. How do you see/think of your nutrition/nutrients/food intake/diet? Do you think what you eat is sufficient? Why?
ii. In your opinion, what makes a diet/meal nutritious/healthy? **[Probe which foods contains required nutrients]**
iii. Would you consider yourself underfed/overfed? Why?
iv. What makes it easy to eat meals/food that contains different nutrients required for adequate nutrition? Why?
v. What are the barriers/makes it difficult to achieve desired nutrition/nutrient intake? Why?
2. Tell me about who makes decisions regarding the foods/meals that are eaten in your household. Why?
3. What influences your decision on the types of foods that you eat? ***[FI: Probe for more details on each mentioned reason below*.**

a. Is it the nutrients the food contains?
b. How do food prices influence what you eat?
c. Do you think your gender influence the food you eat? Why?
d. How does your social status influence your food eating decisions?
e. Seasonality, religion, culture, any other reason.
4. Can you tell me where your foods come from? ***[FI: Probe for Food sources e.g., open market, supermarket, kiosk, etc.]***

a. Do you have a farm? How much of your food that you eat come from come from your farm?
b. Can you tell me why you buy your foods from the places you just mentioned? ***[FI:*** Do you prefer purchasing fresh? How often do you buy food? How does the availability of foods in your area affects your food intake? Are there any other specific factors that influence your choice of food source?

**I would like to ask you a few questions about healthy foods. When we refer to healthy foods, we mean natural foods that are rich in nutrients and could help achieve adequate nutrition such as fruits, vegetables, grains, legumes, roots and tubers, meat, poultry, fish, sea food, eggs, milk.**

5. Do you think you have enough money to buy what you want to eat? What do you think can be done to make it easier for you to be able to get more food intake?
6. Have you heard of highly processed foods such as junks? Can you give me at least, one example of junks?
7. Do you know about the diets/food that contains high sugar, and fats? Can you tell me the problem they can cause to the body?

**I would like to ask you about the influence of the government on healthy food choices.**

8. The government regulates food prices by adding or reducing taxes or by giving assistance/subsidies to farmers. How do these government activities influence your food choices?

**I will now ask you about your health status, activities of daily living, physical function (ability to perform tasks that requires physical strength) affect your food intake/nutrition.**

9. Can you tell me about your health? Can you tell me the health problems you have? Tell me more if your health problems prevent you from the food you eat ***[FI: Probe on health factors e.g., oral health, diabetes, hypertension, muscular/joint pains]*.**
10. Can you tell me how active are you? Do you have issues with your mobility? Do you easily get tired after walking a short distance? Do you have problems with carrying and lifting of utensils in your kitchen when cooking? Do you think you your ability to perform duties that require strength affect how and what you eat?
11. Is there anything else you would like to talk about regarding achieving required adequate nutrition?

We have come to the end of the interview. ***[Thank the participant for participating in the interview]***

